# Built Environment Interventions for Enhancing Youth Engagement in Mental Health and Wellness Activities: A Systematic Review Protocol

**DOI:** 10.1101/2025.10.14.25338032

**Authors:** Ravi Shankar, Jun Wen Joshua, Xu Qian

## Abstract

**Background:** Youth mental health represents a critical public health priority, with built environment interventions increasingly recognized as potential facilitators of engagement in mental health and wellness activities. The physical spaces where young people live, learn, and socialize significantly influence their psychological wellbeing and help-seeking behaviors, yet systematic evidence synthesis on effective built environment strategies remains limited.

**Objective:** To systematically identify, evaluate, and synthesize evidence on built environment interventions that enhance youth engagement in mental health and wellness activities across various settings including educational institutions, community spaces, healthcare facilities, and digital-physical hybrid environments.

**Methods:** This mixed-methods systematic review follows PRISMA-P guidelines and will search PubMed, PsycINFO, Web of Science, Environment Complete, CINAHL, Scopus, and architectural databases from inception to December 2025. Two independent reviewers will screen studies using Covidence software, with no language restrictions applied. Quality assessment will employ the Mixed Methods Appraisal Tool (MMAT) and specialized environmental intervention assessment tools.

**Data Synthesis:** The Socio-Ecological Model will guide synthesis across individual, interpersonal, organizational, community, and policy levels. Quantitative findings will undergo narrative synthesis with effect size reporting where available, while qualitative data will be analyzed through thematic synthesis. Integration will occur through a convergent approach using matrix mapping and joint displays to identify effective design principles and implementation strategies.

## Introduction

The global youth mental health crisis has reached unprecedented levels, with the World Health Organization reporting that one in seven adolescents aged 10-19 experiences a mental health condition, accounting for 15% of the global burden of disease in this age group [1]. Despite the availability of evidence-based interventions, engagement rates among youth remain disappointingly low, with studies indicating that less than 35% of young people with mental health needs receive appropriate services [2, 3]. This engagement gap has prompted exploration of innovative approaches beyond traditional service delivery models, with growing recognition that the built environment—the human-made spaces and places where people live, work, learn, and play—can significantly influence youth mental health outcomes and help-seeking behaviors [4].

The relationship between built environment and mental health operates through multiple pathways including stress reduction through biophilic design elements, social connection facilitation through spatial configuration, identity affirmation through culturally responsive design, and behavioral activation through activity-supporting infrastructure [5, 6]. For youth populations specifically, the built environment holds particular significance as young people spend approximately 87% of their time in indoor environments, primarily within educational institutions, homes, and recreational facilities [7]. These spaces shape not only immediate psychological states but also long-term attitudes toward mental health services and wellness practices, making environmental design a critical yet underutilized tool for mental health promotion [8].

Contemporary understanding of youth engagement in mental health services has evolved from a narrow focus on clinical attendance to encompass broader wellness activities including peer support programs, mindfulness practices, creative therapies, physical activity interventions, and digital mental health tools [9]. This expanded conceptualization recognizes that young people often prefer informal, non-stigmatizing approaches to mental health support that can be integrated into their daily environments and routines. Built environment interventions offer unique opportunities to embed these diverse wellness activities within the spaces youth naturally inhabit, potentially circumventing traditional barriers such as stigma, transportation difficulties, and clinical atmosphere aversion [10].

The theoretical foundations for built environment mental health interventions draw from multiple disciplines including environmental psychology, which elucidates how physical spaces influence cognitive and emotional processes through mechanisms such as attention restoration and stress recovery [11]; architectural phenomenology, which explores how spatial experiences shape consciousness and wellbeing through embodied perception and place attachment [12]; and social ecology, which examines the dynamic interplay between individuals and their multilevel environmental contexts [13]. These theoretical perspectives converge on the understanding that youth mental health cannot be adequately addressed through individual-level interventions alone but requires systemic approaches that modify the environments within which young people develop and seek support.

Recent technological advances have created new possibilities for built environment interventions, including responsive environments that adapt to user needs through sensor-based systems, augmented reality overlays that enhance physical spaces with digital mental health resources, and hybrid spaces that seamlessly integrate virtual and physical wellness activities [14]. The COVID-19 pandemic accelerated experimentation with these innovative approaches, as institutions worldwide rapidly reconfigured physical spaces and developed new environmental strategies to support youth mental health during periods of social distancing and remote learning [15, 16]. These pandemic-driven innovations provide valuable lessons for post-pandemic environmental design, highlighting both opportunities and challenges in creating mental health-promoting spaces for young people.

Despite growing interest in built environment approaches to youth mental health, the evidence base remains fragmented across disciplines including architecture, public health, psychology, education, and urban planning, with limited systematic synthesis of findings. Previous reviews have examined specific aspects such as school design for wellbeing or nature-based interventions for mental health, but comprehensive synthesis examining diverse built environment strategies across multiple settings and their specific impact on youth engagement remains absent [17, 18]. This gap limits the ability of designers, policymakers, and mental health professionals to make evidence-informed decisions about environmental investments and interventions.

### Theoretical Framework

This systematic review employs the Socio-Ecological Model (SEM) as its primary organizing framework, recognizing that youth engagement in mental health and wellness activities results from complex interactions between individual characteristics and multilevel environmental influences [19]. The SEM provides a comprehensive structure for categorizing and analyzing built environment interventions across five nested levels: individual, interpersonal, organizational, community, and policy, each offering distinct opportunities for environmental modification to enhance youth mental health engagement.

At the individual level, built environment interventions influence personal factors including sensory processing, emotional regulation, cognitive restoration, and identity expression [20]. Environmental features such as lighting quality, acoustic conditions, color schemes, and spatial proportions directly impact neurophysiological stress responses and attention capacity, particularly important for youth experiencing anxiety, depression, or neurodevelopmental conditions. The framework recognizes that individual responses to environmental features vary based on developmental stage, cultural background, previous trauma exposure, and sensory sensitivities, necessitating flexible and inclusive design approaches that accommodate diverse youth populations.

The interpersonal level examines how built environments facilitate or hinder social connections crucial for youth mental health, including peer relationships, family involvement, and therapeutic alliances [21]. Spatial configurations influence social dynamics through factors such as proximity, privacy gradients, territorial definition, and activity affordances that shape interaction patterns. For youth mental health engagement, environments must balance opportunities for social connection with provisions for solitude and emotional regulation, recognizing that both social support and personal space contribute to wellbeing.

At the organizational level, the framework considers how institutional environments— schools, clinics, community centers, residential facilities—structure youth mental health engagement through spatial organization, resource allocation, and activity programming [22]. Built environment features at this level include wayfinding systems that reduce navigation stress, flexible spaces that accommodate diverse wellness activities, and design elements that communicate institutional values regarding mental health. The framework emphasizes how organizational environments can either reinforce or challenge mental health stigma through design choices that normalize or marginalize wellness activities.

The community level encompasses broader neighborhood and municipal environments that influence youth access to mental health resources and wellness opportunities [23]. This includes the spatial distribution of mental health services, transportation infrastructure connecting youth to resources, public spaces supporting wellness activities, and neighborhood features influencing safety perceptions and stress exposure. The framework recognizes that community-level environmental factors often represent the most significant barriers to mental health engagement for marginalized youth populations.

The policy level examines how regulations, standards, and guidelines shape built environment interventions for youth mental health across all other levels [24]. This includes building codes affecting therapeutic space design, educational facility standards influencing wellness resource allocation, zoning regulations determining service accessibility, and funding mechanisms supporting environmental modifications. The framework emphasizes that sustainable built environment interventions require supportive policy contexts that recognize environmental design as a legitimate mental health intervention strategy.

### Objectives

This systematic review aims to comprehensively synthesize evidence on built environment interventions designed to enhance youth engagement in mental health and wellness activities across diverse settings and populations. The primary objective encompasses identifying effective environmental design strategies, understanding implementation processes, and evaluating outcomes related to youth mental health service utilization, wellness activity participation, and psychosocial wellbeing indicators. Specific objectives include: (1) categorizing built environment interventions by design features, settings, and theoretical foundations while examining their differential impacts on youth engagement outcomes; (2) identifying barriers and facilitators to implementing built environment interventions within various institutional and community contexts; (3) examining how intervention effectiveness varies across youth subpopulations defined by age, cultural background, socioeconomic status, and mental health needs; and (4) developing an evidence-based framework linking specific environmental design principles to youth engagement mechanisms and outcomes.

## Methods

### Study Design

This systematic review protocol follows the Preferred Reporting Items for Systematic Review and Meta-Analysis Protocols (PRISMA-P) guidelines and employs a convergent mixed-methods design to synthesize quantitative and qualitative evidence on built environment interventions for youth mental health engagement [25]. The mixed-methods approach recognizes that understanding environmental interventions requires both outcome evaluation and process understanding, integrating effectiveness data with implementation experiences and contextual factors. The review will be registered with PROSPERO upon finalization of the review team.

### Search Strategy

A comprehensive search strategy developed with a health sciences librarian will be implemented across multiple databases from inception to December 2025. Primary databases include PubMed/MEDLINE, PsycINFO, Web of Science Core Collection, Environment Complete, CINAHL, and Scopus. Specialized databases including Avery Index to Architectural Periodicals, Design and Applied Arts Index, and Urban Studies Abstracts will capture architecture and planning literature. Grey literature searches will encompass governmental reports, organizational websites, and dissertation databases.

The search strategy combines four concept clusters using Boolean operators: (1) Youth terms including “adolescent*,” “youth,” “young people,” “teen*,” “emerging adult*”; (2) Mental health and wellness terms including “mentalhealth,””wellbeing,” “psychological,””wellness,” “resilience,” “mindfulness,” “therapy,” “counseling”; (3) Engagement terms including “engagement,” “participation,” “utilization,” “access,” “uptake,” “involvement,” “help-seeking”; (4) Built environment terms including “built environment,” “physical environment,””space design,” “architecture,” “environmental design,” “healing environment,” “therapeutic landscape,” “biophilic design.” No language or date restrictions will be applied to ensure comprehensive coverage.

### Eligibility Criteria

Studies will be included if they: (1) focus on youth populations aged 10-25 years; (2) evaluate built environment interventions designed to influence mental health or wellness engagement; (3) report engagement-related outcomes including service utilization, activity participation, help-seeking behaviors, or related psychosocial measures; (4) employ empirical research designs including experimental, quasi-experimental, observational, or qualitative methodologies. Mixed-age studies will be included if youth-specific data are reported separately.

Exclusion criteria include: (1) studies focusing exclusively on clinical treatment outcomes without engagement measures; (2) natural environment interventions without built environment components; (3) virtual reality interventions without physical environment integration; (4) commentaries, editorials, or papers without empirical data; (5) studies where environmental modification represents an incidental rather than intentional intervention component.

### Study Selection

Study selection will proceed through two stages using Covidence systematic review software. Two independent reviewers will screen titles and abstracts against eligibility criteria, with conflicts resolved through discussion or third reviewer consultation. Full-text screening will follow the same dual-review process with documented reasons for exclusion. A PRISMA flow diagram will track study selection processes. Inter-rater reliability will be calculated using Cohen’s kappa, with values _≥_0.75 considered acceptable.

### Data Extraction

A standardized extraction form developed and piloted within Covidence will capture: study characteristics (authors, year, country, setting, design); participant characteristics (age range, gender distribution, mental health status, socioeconomic indicators); intervention details (environmental modifications, theoretical basis, implementation process, duration); engagement outcomes (measures used, time points, effect sizes); implementation factors (costs, stakeholder involvement, sustainability); and contextual information (cultural considerations, policy environment, COVID-19 adaptations).

For qualitative studies, extraction will include methodological orientation, data collection methods, analytical approaches, key themes, and illustrative quotes. The Socio-Ecological Model will guide categorization of interventions and outcomes across system levels. Extraction will be completed independently by two reviewers with discrepancy resolution through consensus discussion.

### Quality Assessment

Methodological quality will be assessed using the Mixed Methods Appraisal Tool (MMAT) version 2018, enabling consistent evaluation across diverse study designs [26]. Additional assessment will employ the Quality Assessment Tool for Quantitative Studies for intervention studies and the CASP Qualitative Checklist for qualitative research [27]. Environmental intervention-specific quality criteria will include: clarity of environmental modification description, fidelity assessment, contamination control, and setting characteristic reporting.

Two reviewers will independently conduct quality assessments with disagreements resolved through discussion. Quality scores will not determine study exclusion but will inform sensitivity analyses and confidence in findings assessment using GRADE-CERQual for qualitative evidence and standard GRADE for quantitative outcomes [28].

### Data Synthesis

Quantitative synthesis will employ narrative approaches due to anticipated heterogeneity in interventions, settings, and outcomes. Where feasible, effect sizes will be calculated and reported with 95% confidence intervals. Harvest plots will visualize patterns of effectiveness across intervention types and outcomes. Subgroup analyses will explore variations by age group, setting type, intervention intensity, and implementation quality.

Qualitative synthesis will follow Thomas and Harden’s thematic synthesis approach involving line-by-line coding, descriptive theme development, and analytical theme generation [29]. The Socio-Ecological Model will provide an organizing framework while allowing emergent themes beyond predetermined categories. NVivo software will facilitate coding and theme development with regular team meetings ensuring coding consistency.

Integration of quantitative and qualitative findings will follow Fetters et al.’s convergent approach using joint displays and narrative weaving [30]. A matrix will map intervention components to outcomes and implementation factors, identifying patterns of convergence, complementarity, and divergence between evidence types. The integrated synthesis will generate a conceptual model linking environmental design principles to engagement mechanisms and outcomes.

## Discussion

This systematic review will provide the first comprehensive synthesis of built environment interventions for enhancing youth engagement in mental health and wellness activities, addressing a critical gap in understanding how physical spaces can be leveraged to improve youth mental health outcomes. The findings will have important implications for multiple stakeholders including architects and designers who shape youth-serving spaces, mental health professionals seeking to enhance service engagement, educators and administrators managing institutional environments, policymakers allocating resources for mental health infrastructure, and young people themselves who navigate these environments while seeking support.

The theoretical contributions of this review include advancing understanding of environment-behavior relationships in youth mental health contexts, extending the Socio-Ecological Model’s application to built environment interventions, and identifying mechanisms through which environmental modifications influence help-seeking behaviors and wellness participation. The review will contribute to emerging transdisciplinary frameworks that bridge environmental design, public health, and youth development fields, potentially catalyzing new collaborative approaches to mental health promotion.

Practical implications encompass evidence-based design guidelines for youth mental health facilities, schools, and community spaces that prioritize engagement over purely clinical considerations. The review will identify cost-effective environmental modifications that can be implemented within existing budget constraints, particularly important for resource-limited settings serving marginalized youth populations. By highlighting successful implementation strategies and common pitfalls, the findings will support organizations planning environmental interventions to enhance youth mental health engagement.

The review will likely reveal significant evidence gaps requiring future research attention, including limited longitudinal studies examining sustained engagement impacts, insufficient attention to cultural variations in environmental preferences and responses, lack of economic evaluations comparing built environment interventions to traditional engagement strategies, and minimal investigation of digital-physical hybrid environments emerging post-pandemic. These gaps will inform research funding priorities and dissertation topics for emerging scholars interested in youth mental health and environmental design intersections.

Methodological strengths include the comprehensive search strategy spanning multiple disciplines, use of established frameworks guiding systematic synthesis, inclusion of diverse evidence types capturing intervention complexity, and transparent reporting following PRISMA-P guidelines. The mixed-methods approach enables nuanced understanding beyond simple effectiveness evaluations, while the Socio-Ecological Model ensures attention to multilevel influences often overlooked in individual-focused mental health research.

Limitations include potential publication bias toward positive findings, as unsuccessful interventions may remain unpublished. Language restrictions during full-text review may exclude relevant non-English studies, particularly from countries with different architectural traditions and mental health approaches. The broad age range (10-25 years) encompasses diverse developmental stages with potentially different environmental needs and preferences. Heterogeneity in intervention types, settings, and outcomes may preclude meta-analysis, limiting quantitative synthesis precision.

The review’s findings will be disseminated through peer-reviewed publications, conference presentations targeting both mental health and design communities, policy briefs for governmental and organizational decision-makers, and accessible summaries for youth advocacy groups. A dedicated website will provide open access to the review protocol, findings, and supplementary materials including the conceptual framework and design recommendations. Social media campaigns will engage young people in discussing how their environments influence mental health engagement.

This systematic review represents a crucial step toward recognizing built environment design as a legitimate and powerful strategy for addressing the youth mental health crisis. By synthesizing evidence across disciplines and settings, the review will provide actionable insights for creating spaces that not only accommodate but actively promote youth engagement in mental health and wellness activities. The ultimate goal is contributing to a future where all young people have access to environments that support their mental health journey, reducing barriers to help-seeking and normalizing wellness practices through thoughtful design.

Future implications include potential development of environmental assessment tools specifically evaluating youth mental health engagement potential, integration of evidence-based design principles into architectural education curricula, and establishment of certification programs for youth mental health-promoting environments similar to existing sustainability certifications. The review may catalyze policy reforms mandating mental health considerations in youth facility design standards and funding mechanisms supporting environmental interventions as preventive mental health investments.

## Data Availability

All data produced in the present work are contained in the manuscript

## Notes

### Competing Interest Statement

The authors have declared no competing interest.

### Funding Statement

This study did not receive any funding

